# Modelling the positive testing rate of COVID-19 in South Africa Using A Semi-Parametric Smoother for Binomial Data

**DOI:** 10.1101/2020.11.11.20230250

**Authors:** Olajumoke Evangelina Owokotomo, Samuel Manda, Adetayo Kasim, Jurgen Claesen, Ziv Shkedy, Tarylee Reddy

## Abstract

The current outbreak of COVID-19 is a major pandemic that has shaken up the entire world in a short time. South Africa has the highest number of COVID-19 cases in Africa and understanding the country’s disease trajectory is important for government policy makers to plan the optimal COVID-19 intervention strategy. The number of cases is highly correlated with the number of COVID-19 tests undertaking. Thus, current methods of understanding the COVID-19 transmission process in the country based only on the number of cases can be misleading. In light of this, we propose to estimate both the probability of positive cases per tests conducted (the positive testing rate) and the rate in which the positive testing rate changes over time (its derivative) using a flexible semi-parametric model.

We applied the method to the observed positive testing rate in South Africa with data obtained from March 5th to September 2nd 2020. We found that the positive testing rate was declining from early March when the disease was first observed until early May where it kept on increasing. In the month of July 2020, the infection reached its peak then its started to decrease again indicating that the intervention strategy is effective. From mid August, 2020, the rate of change of the positive testing rate indicates that decline in the positive testing rate is slowing down, suggesting that a less effective intervention is currently implemented.

## 1 Introduction

Coronaviruses are a large family of viruses which may cause respiratory infections ranging from the common cold to more severe diseases such as Middle East Respiratory Syndrome (MERS) and Severe Acute Respiratory Syndrome (SARS). The ongoing outbreak of the novel coronavirus (SARS-CoV-2) was first detected on 31st December 2019 in Wuhan, China (World Health Oragnisation, 2020). The virus has rapidly spread with a total of 37,423,660 confirmed cases and 1,074,817 deaths as of 12th October 2020 (World Health Oragnisation, 2020).

The first COVID-19 case in South Africa was reported on March 5th 2020. By October 12th, 2020, South Africa had the highest burden of COVID-19 cases in the African region with 692,471 reported cases and 17,780 confirmed COVID-19 related deaths(World Health Oragnisation, 2020). The South African government declared a national state of disaster on March 15th 2020 and commenced a state of lockdown from March 26th 2020 in an effort to reduce COVID-19 transmission in the country(Reddy et al., 2020). During this period all international and inter-provincial borders were closed, as well as the education sector and several economic sectors in the country. As of June 2020, the country adopted a COVID-19 risk-adjusted strategy with a phased re-opening of selected economic sectors and schools.

Modeling the number of COVID-19 cases and in particular producing a reliable short and long term prediction for the number of COVID-19 cases become a central tool for policy makers to design innervation strategies in order to control the disease’s spread. Recently, (Reddy et al., 2020) proposed a robust model based approach, that does not require to make assumptions about the transmission process to model the number of COVID-19 cases and to provide a short term prediction for 5-10 days ahead. These non-linear epidemiological models have previously been applied to model other disease outbreaks such as Ebola (Chowell et al., 2019), Dengue (Hsieh and Chen, 2009), Zika virus (Sebrango-Rodríguez et al., 2017) and, more recently, the COVID-19 pandemic (Roosa et al., 2020); (Shen, 2020); (Tariq et al., 2020). Specifically, Roosa et al. (2020) fitted the generalized logistic model, Richards’s model and a sub-epidemic model to the cumulative COVID-19 cases in the Hubei province of China and produced a short-term forecast of 5, 10 and 15 days ahead .The authors also expanded on this work for the province of Guandong. In the recent analysis by Shen (2020), a similar approach was taken to estimate the key epidemic parameters for all 11 provinces in China as well as 9 selected countries.

All models discussed above made use of the daily or cumulative number of cases to estimate the models and the parameters of interest.In the context of COVID-19, this introduces a difficulty as seen in Figure 1, since in South Africa (and many other countries) the number of tests and number of cases are correlated (Reddy et al., 2020).

**Figure 1:**
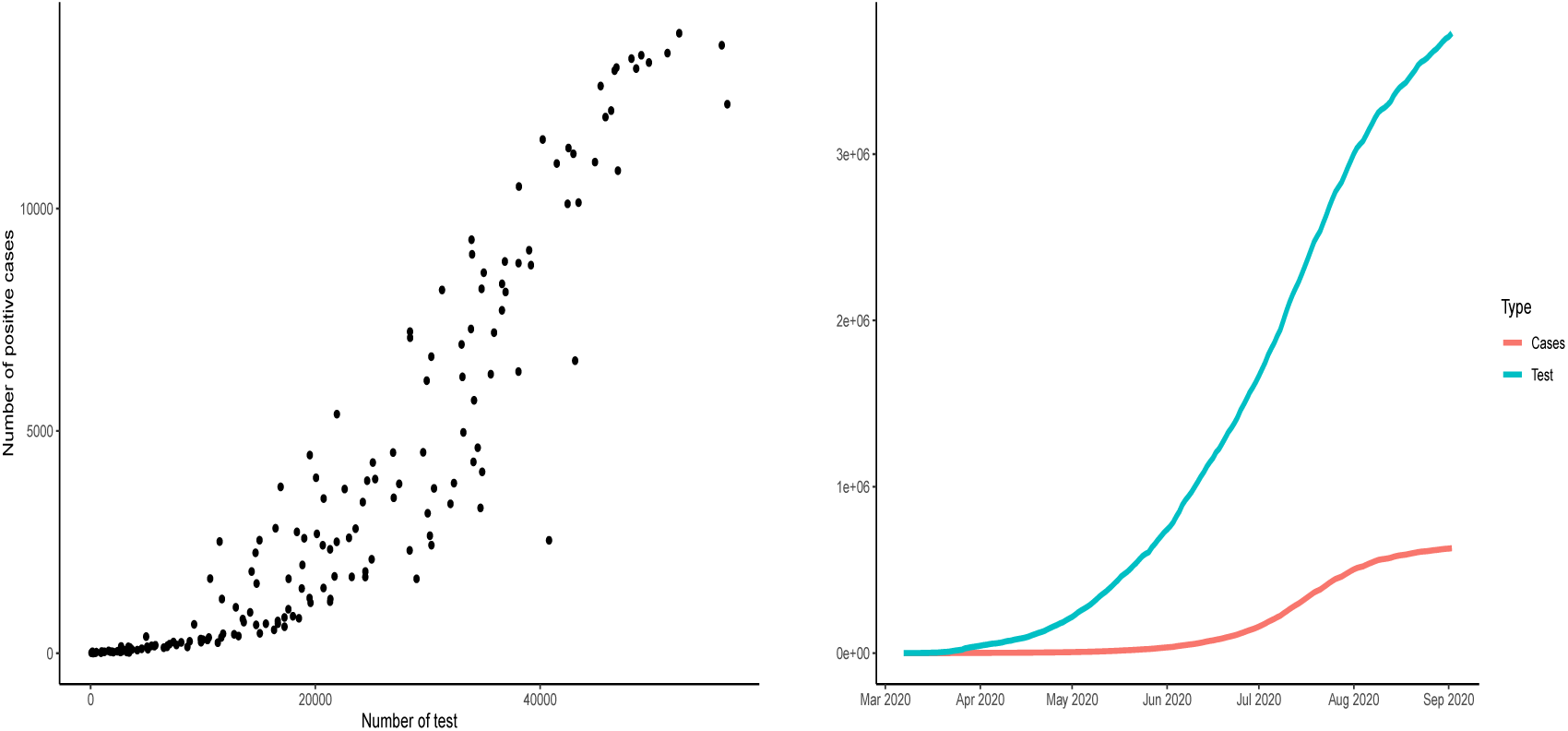
Panel a: Relationship between the daily number of tests and the daily number of positive cases. Panel b:The total number of cases and total number of COVID-19 tests carried out between the period March 7th 2020 and September 2nd 2020. Spearman’s rank correlation between the number of tests to the number of cases is equal to 0.9632752, (p-value < 2.2e-16).

Positive testing rate, i.e., the probability of positive case among the total number of tests, has been seen as an important metrics in understanding the transmission of COVID-19 in the literature (Our world in Data, 2020). Due to the correlation (dependence) of the number of cases on the number of tests conducted, no country knows the actual total number of people infected with COVID-19 but only the infection status of those who have been tested. Therefore, in countries with a high positive rate, the number of confirmed cases is more likely to represent only a small proportion of the true number of cases. However, when the positive rate increases it can suggest the virus is actually spreading faster than the growth seen in confirmed cases. On May 12, 2020 the World Health Organization (WHO) advised governments that before relaxing intervention measures, rates of positively in testing should remain at 5% or lower for at least 14 days (John Hopkins coronavirus resource center,2020, WHO, 2020).

To overcome the problem that the number of cases depends on the number of tests, we propose an alternative modeling approach that focuses on COVID-19 positive testing rate, i.e., the probability of positive cases per tests conducted. In this paper we model the daily number of COVID-19 cases among the number of tests carried out using a semi-parametric model in which the rate of change of the positive testing rate is estimated using a smooth function of time. In particular, we apply scatterplot smoothing techniques for binomial data using generalized additive models in order to obtain an estimate of the rate of change Ruppert et al. (2003). In Section 2 we describe the testing policy in South Africa from which the data used for the analysis presented in this paper was obtained. The modelling approach, the model formulation for the positive testing rate and the methodology to construct simaultenous confidence bands are discussed in Section 3. Section 4 contains the results, and the discussions and conclusions are in Section 5.

## 2 Data

### 2.1 Daily number of tests and confirmed cases

The daily number of reported COVID-19 cases and tests for the period of March 7th 2020 to September 2nd 2020 is presented in Figure 2. The growth of COVID-19 infections in South Africa appears to be tri-phasic especially during the early phase when the cumulative cases were low with rapid growth until March 27th 2020. A total of 243 daily new cases were observed, followed by a sharp decline in the rate of new cases. From March 28th 2020 to April 6th 2020 the daily increase in cases was consistently below 100. From May 2020 onwards, a consistent increase of more than 1000 cases per day were observed. The peak period was between of July 9th and 19th where more than 10,000 reported cases were reported on a daily basis. As of July, a total of 3726721 tests had been conducted, corresponding to a testing rate of 22.816 per 1000 population. Throughout this period, the proportion of infections increased until mid July when it started to decrease.

**Figure 2:**
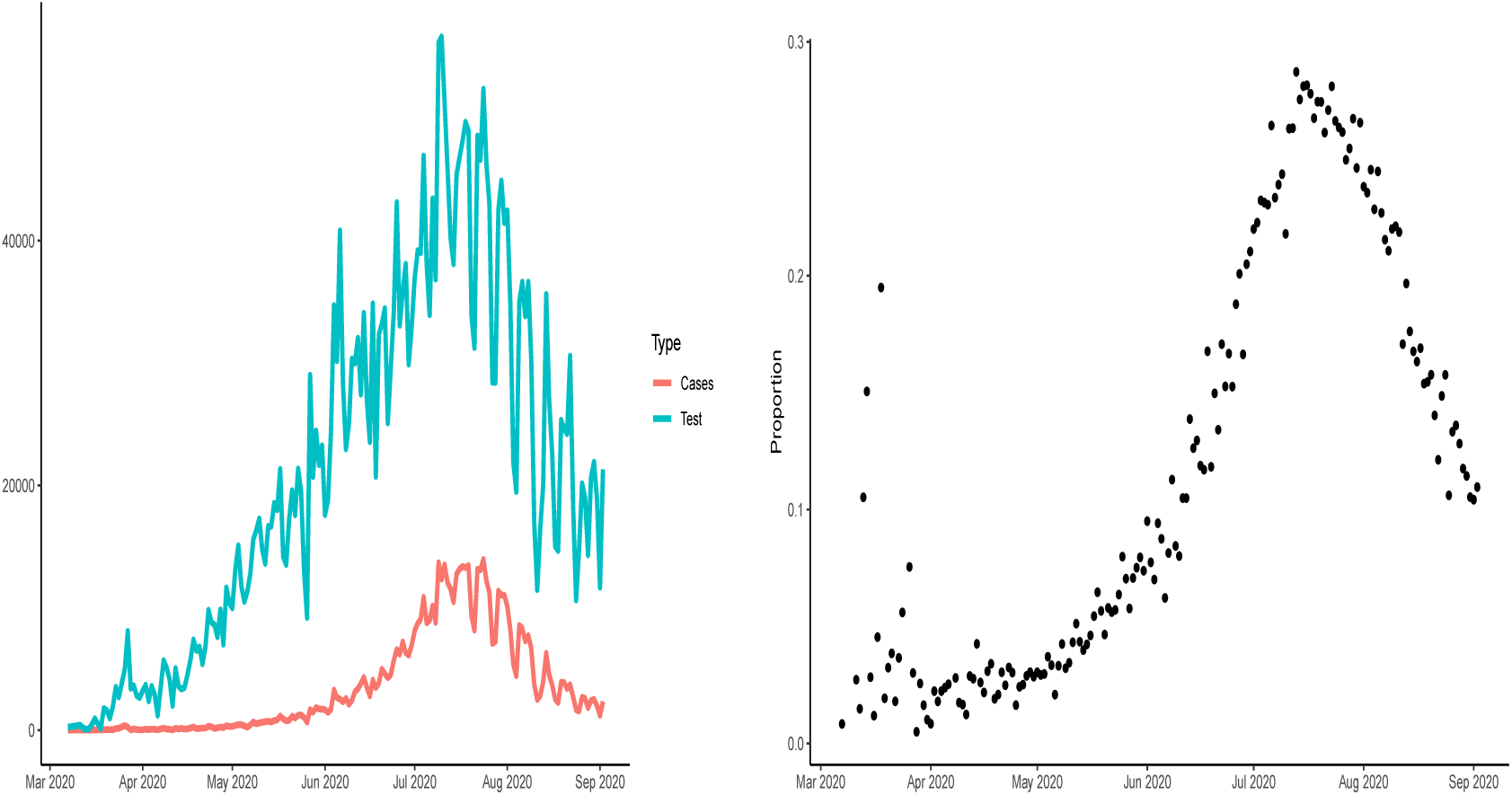
Panel a: Daily number of cases and daily number of COVID-19 tests between March 7th, 2020 and September 2nd, 2020. Panel b: Positive testing rate.

### 2.2 Testing policy in South Africa

A total of 3,245,087 tests for SARS-COV-2 were conducted between March 1st and August 29th 2020. These tests were performed on individuals who satisfied the case definition for persons under investigation (PUI). The PUI definition, which was amended consistently included at least one of the following criteria: symptomatic individuals seeking testing, hospitalized individuals for whom testing was done, individuals in high-risk occupations (e.g health care workers), individuals in outbreak settings, and individuals identified through community screening and testing programmes which were implemented between April and the middle of May 2020. The number of tests performed on a weekly basis increased from March 2020 until the third week of May and proceeded to decrease over the subsequent two weeks due to a limited supply of testing kits. The average time elapsed from specimen collection to testing was under two days in both the private and public sectors from August 22th to 29th August 2020.

## 3 Modeling COVID-19 infection rate in South Africa using Generalized Linear Mixed Effects Model for Binary Data

### 3.1 Model formulation for the positive testing rate

The number of positive cases is assumed to be binomially distributed. Let *π*_*t*_ be the daily positive testing rate per test, *Y*_*t*_ be the daily number of cases and *n*_*t*_ be the daily number of tests. Our aim is to model the probability *π*_*t*_ and to produce a model-based estimate for its first derivative, i.e., the change in the positive testing rate over time. Semi-parametric regression model for binomial data was used to provide an estimate of the positive testing rate as a function of time. The relationship can be expressed as

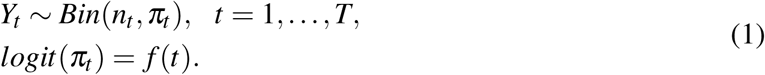

Here, *f* (*t*) is a smooth function of the time *t*. Smoothing splines are commonly used for this purpose (Ruppert et al. (2003)). A general spline model of degree *d* with *K* knots can be written as follows:

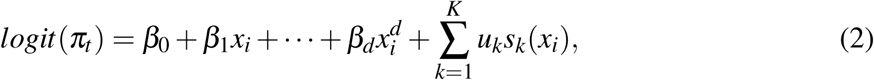

where *s*_*k*_(*x*) is a set of spline basis functions.

To avoid overfitting, the spline model is typically estimated by considering penalized maximum likelihood estimation, with a penalty term of the form 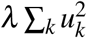. Ruppert et al. (2003) showed that the penalized regression problem can be expressed as an equivalent generalized linear mixed-effects model (GLMM):

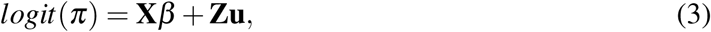

with *π* = [*π*_1_, *π*_2_, …, *π*_*T*_]^*T*^, *β* = [*β*_0_, *β*_1_, …, *β*_*d*_]^*T*^, and **u** = [*u*_1_, *u*_2_, …, *u*_*K*_]^*T*^. Note that *β* and **u** are vectors of the fixed and random effects, respectively, with 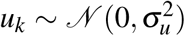 where 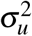 acts as the smoothing parameter. This representation has the advantage that the degree of smoothing can be estimated from the data using standard mixed-model software (e.g., Ruppert et al. (2003), chapter 4). The design matrices **X** and **Z** are defined as follows:

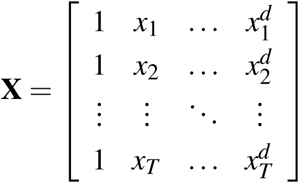

and

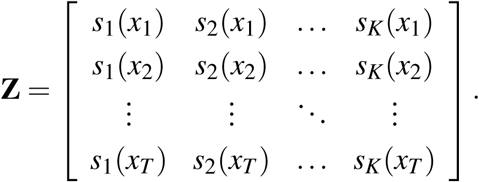

The estimation of the model (3) is performed by means of penalized quasi-likelihood (PQL). Initial estimates for *β* and **u** are used to calculate the pseudo-data **y**\*:

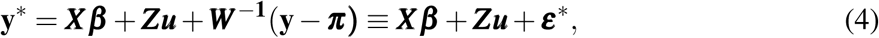

where ***W*** is a diagonal matrix with variances of *y*_*i*_ on the diagonal. The pseudo-error ***ε***\* has a variance-covariance matrix ***R*** = ***W*** ^−1^*ϕ*, where *ϕ* is the dispersion parameter, equal to one for the standard binomial model family. Equation (4) resembles a linear mixed-effects model (LMM) formulation for **y**\*. Thus, an LMM is fitted to the pseudo-data, yielding updated estimates of ***β***, ***u***, 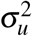, and *ϕ*. The procedure of calculating pseudo-data and re-fitting the LMM is repeated until convergence.

### 3.2 Estimating the derivative for *π*_*t*_

Once the positive testing rate, *π*_*t*_, is estimated according to Equation (1) we can estimate the rate of change in the positive testing rate over time using the derivative of *π*_*t*_ given by

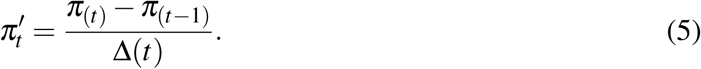

Note that if the number of tests is constant over time and applied to a random sample of the population, 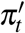 can give an indication to the change in the virus’ transmission in the population (since in this case, it is gives the change in transmission probability). However, it is unlikely to assume that the number of tests will be constant nor that the tests will be applied to random sample from the population. Also in this case, the derivative can provides a good indication about the general trend of the virus’ transmission for the tested population and can be used as a tool to asses the success of an implemented intervention strategy.

### 3.3 Construction of pointwise confidence band

According to Ruppert et al. (2003), an approximate 100(1-*α*)% pointwise confidence band for an estimated penalized spline in the GLMM framework, 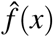, is given by:

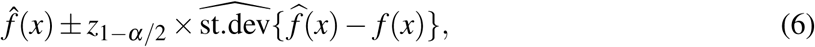

where

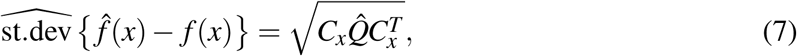

with *C*_*x*_ =(1 *x* … *x*^*d*^ *s*_1_ (*x*) … *s*_*K*_(*x*)) and

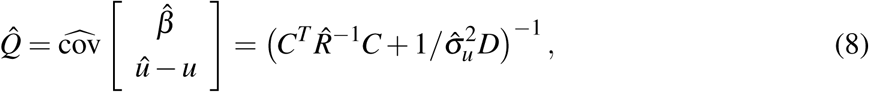

where *C* = [*XZ*] and 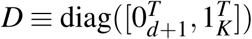

Pointwise confidence bands, however, need to be corrected for multiplicity. Also, they ignore serial correlation. Therefore, we make use of simultaneous confidence bands implemented in Claesen1 et al. (2013), which allow to make joint statements on multiple locations of the fitted curve. A 100(1-*α*)% simultaneous confidence band for 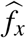 is defined as:

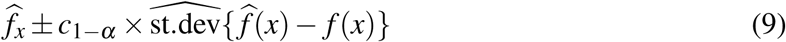

where the critical value, *c*_1−*α*_, is the (1-*α*) quantile of the random variable

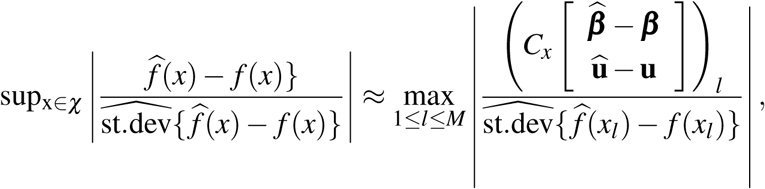

which can be found by simulating from an approximate multivariate normal distribution

## 4 Application to the data

A generalized additive model was fitted to the data with the time component was used as the smooth term.The model was fitted using the `gam()` function of the `mgcv` (Wood, 2017) library in R (R Core Team, 2020). Figure 3,shows that the estimated positive testing rate peaked on July 19th, 2020, at the same time that the number of tests was at its highest level. From that time onward, both number of tests and the positive testing rate declined. This could be a result of a reduction of the virus’ transmission in the population or a result of a change in the population to which the tests were applied.

**Figure 3:**
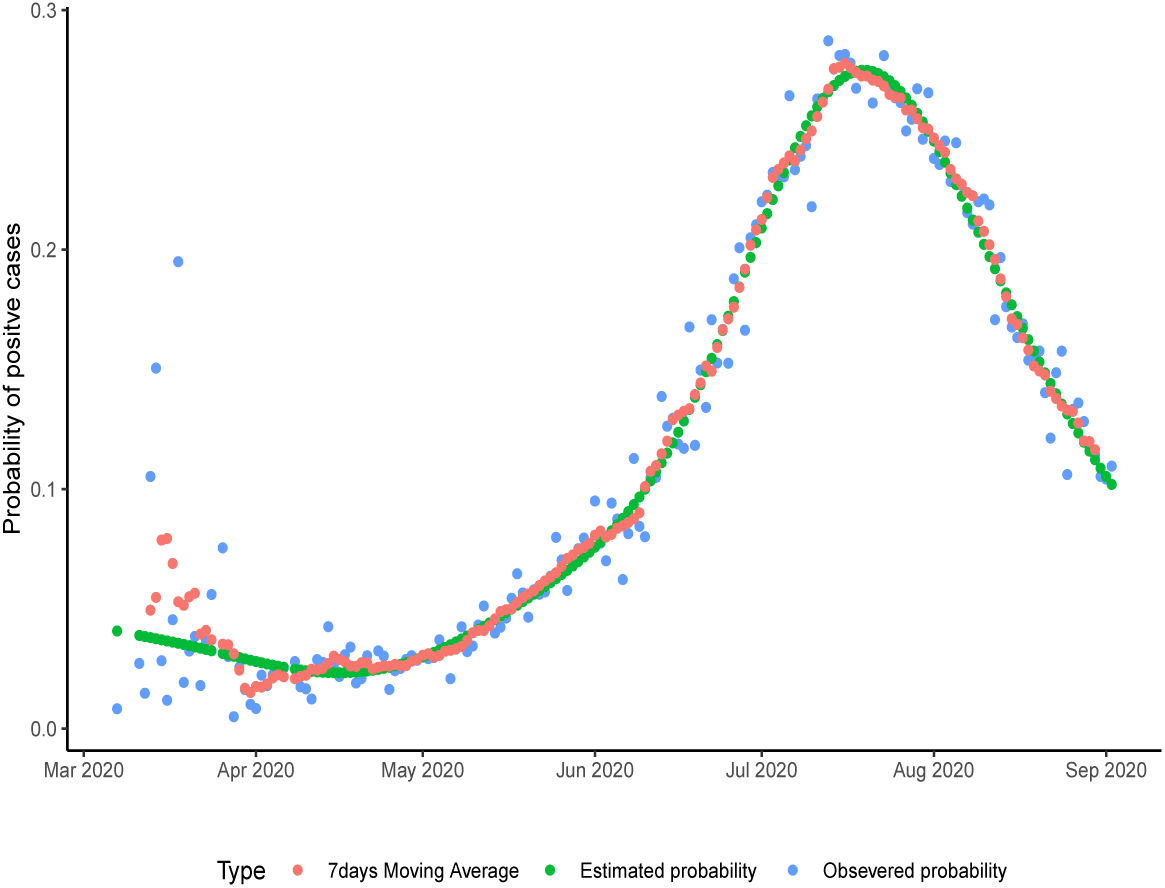
Observed proportion of infection over time, the estimated probability and the 7 days moving average of positive testing rate.

A commonly used measure in understanding COVID-19 transmission rate is the moving average. A 7 days moving average for the positive testing rate was also estimated and this gave a similar evolution pattern as the estimated positive testing rate.

From July 19 onward, the change in positive testing rate (the derivative plot presented in upper part of Figure 4) is negative (indicating decline in the positive testing rate) but from August, 11, 2020, the derivative begins to increase. This could suggest a change in the transmission trend and an increasing number of positive cases.

**Figure 4:**
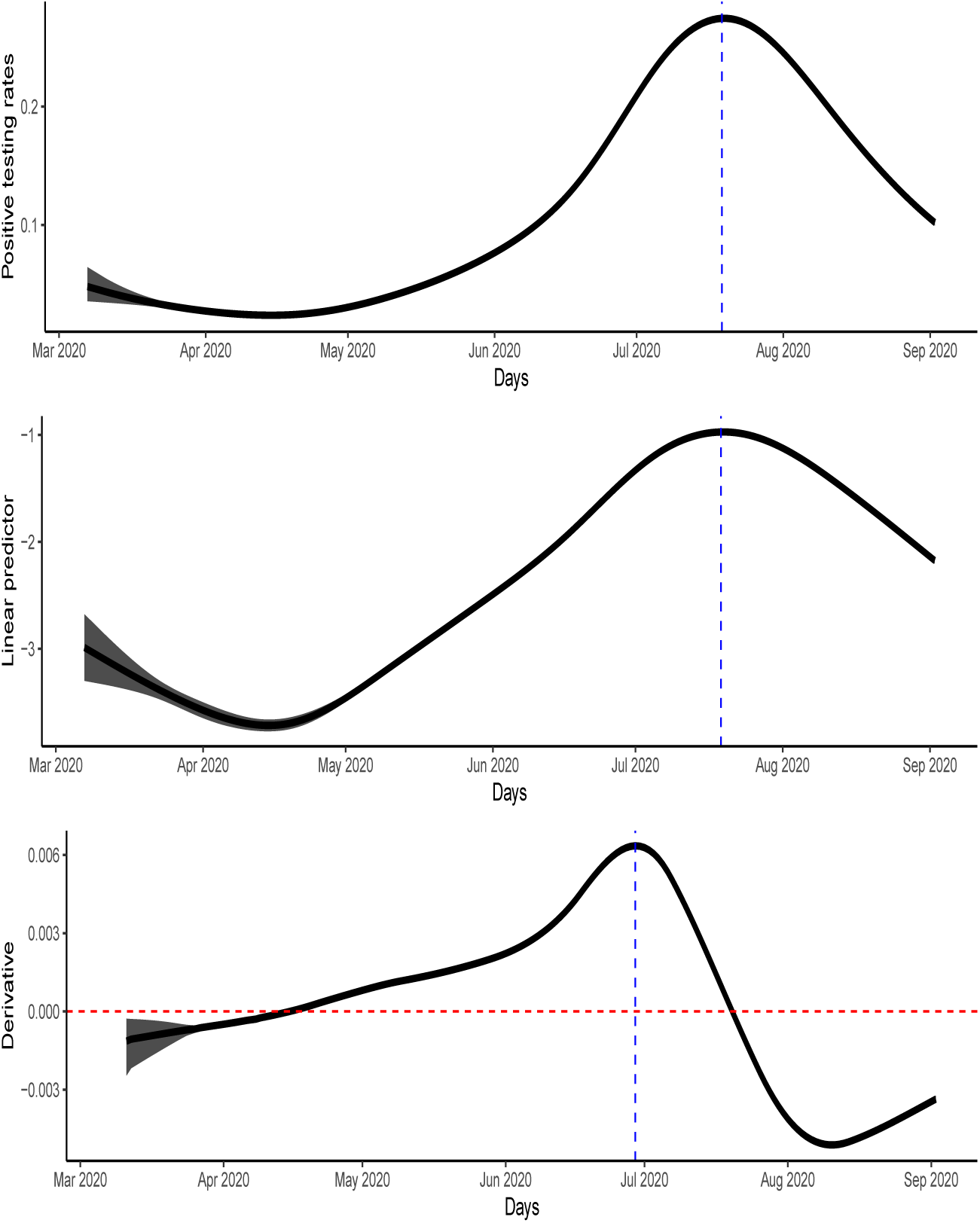
Upper panel: Estimated positive testing rate with 95% simultaneous confidence band. Middle panel: The linear predictor of the smoother with 95% simultaneous confidence band. Lower panel: The derivative of the estimated probability with 95% simultaneous confidence band.

## 5 Discussion

In view of the existing healthcare challenges in South Africa, reliable and accurate knowledge about the positive testing rate of COVID-19 is important to ensure prediction of the disease trajectory, optimal resource allocation and better understanding of the transmission process. In the current study we modelled the COVID 19 cases out of the number of tests as a function of time using semi-parametric approach. This approach allows us to adjust for or take into account the number of tests performed, which when ignored may lead to erroneous conclusions. Also, this method allows us to overcome the problem to modelling the number of cases alone and to take into account the strong relationship between the number of cases and the number of tests which can lead to a misleading result and therefore affect government policy regarding measures and precautions needed.

The positive testing rate decreased from early March when the disease was first observed until early May when it kept on increasing. In July, the infection reached its peak and then consistently decreased, indicating that the intervention strategy was effective. From mid August, 2020, the rate of change of the positive testing rate indicates the decline in the positive testing rate is slowing down suggesting that a less effective intervention is currently implemented. The moving average is another measure that can be used to understand the rate of infection, but unlike the positive testing rate, the moving average commonly uses partial information since there is always loss of information on both tails. In our case, the same result was obtained using both measures.

The rate of infection can be used as an indicator for the evolution of the outbreak over time and to reveal new trends in the outbreak. One could also extend our approach by modeling jointly the number of tests and number of positive cases. These results need to be interpreted under the background of changing COVID-19 testing strategies in the country. When the positive testing rate is tracked in real time, it can provide useful guidance to policy makers

## Data Availability

data are publicly available

